# Development and validation of a neural network-based survival model for mortality in ischemic heart disease

**DOI:** 10.1101/2023.06.16.23291527

**Authors:** Peter C. Holm, Amalie D. Haue, David Westergaard, Timo Röder, Karina Banasik, Vinicius Tragante, Alex H. Christensen, Laurent Thomas, Therese H. Nøst, Anne-Heidi Skogholt, Kasper K. Iversen, Frants Pedersen, Dan E. Høfsten, Ole B. Pedersen, Sisse Rye Ostrowski, Henrik Ullum, Mette N. Svendsen, Iben M. Gjødsbøl, Thorarinn Gudnason, Daníel F. Guðbjartsson, Anna Helgadottir, Kristian Hveem, Lars V. Køber, Hilma Holm, Kari Stefansson, Søren Brunak, Henning Bundgaard

## Abstract

**Background:** Current risk prediction models for ischemic heart disease (IHD) use a limited set of established risk factors and are based on classical statistical techniques. Using machine-learning techniques and including a broader panel of features from electronic health records (EHRs) may improve prognostication.

**Objectives:** Developing and externally validating a neural network-based time-to-event model (PMHnet) for prediction of all-cause mortality in IHD.

**Methods:** We included 39,746 patients (training: 34,746, test: 5,000) with IHD from the Eastern Danish Heart Registry, who underwent coronary angiography (CAG) between 2006-2016. Clinical and genetic features were extracted from national registries, EHRs, and biobanks. The feature-selection process identified 584 features, including prior diagnosis and procedure codes, laboratory test results, and clinical measurements. Model performance was evaluated using time-dependent AUC (tdAUC) and the Brier score. PMHnet was benchmarked against GRACE Risk Score 2.0 (GRACE2.0), and externally validated using data from Iceland (n=8,287). Feature importance and model explainability were assessed using SHAP analysis.

**Findings:** On the test set, the tdAUC was 0.88 (95% CI 0.86-0.90, case count, cc=196) at six months, 0.88(0.86-0.90, cc=261) at one year, 0.84(0.82-0.86, cc=395) at three years, and 0.82(0.80-0.84, cc=763) at five years. On the same data, GRACE2.0 had a lower performance: 0.77 (0.73-0.80) at six months, 0.77(0.74-0.80) at one year, and 0.73(0.70-0.75) at three years. PMHnet showed similar performance in the Icelandic data.

**Conclusion:** PMHnet significantly improved survival prediction in patients with IHD compared to GRACE2.0. Our findings support the use of deep phenotypic data as precision medicine tools in modern healthcare systems.

## Introduction

In patients with ischemic heart disease (IHD), improved clinical application of the wide array of prognostic risk factors and disease markers may inform treatment options for the individual patient^1–3^. For example, the updated version of Global Registry of Acute Coronary Events (GRACE) score (GRACE2.0) received a class IIa recommendation for assessing risk and management of patients with non–ST-elevation myocardial infarction (nSTEMI) in the 2020 European Society of Cardiology (ESC) guidelines^1^. However, GRACE2.0 and other traditional risk scoring schemes in IHD such as Framingham and Thrombolysis in Myocardial Infarction (TIMI) use a limited set of input features (<10) and likely underutilize most data available in modern electronic health records (EHRs)^4–7^.

Integrating a richer set of input features could be overcome with machine learning (ML) models such as neural networks. These can capture non-linear interactions without the need for imputation of missing data or expert feature engineering^8, 9^, leveraging the multitude of heterogeneous healthcare data stored in modern EHRs and national registries in the development of clinical decision support tools.

ML-based approaches have shown promising results for risk-estimation in cardiology with better performance than traditional models^10–13^. In patients with stable IHD, Motwani et al. showed that an ML algorithm combining clinical variables with imaging variables from coronary CT angiography predicted 5-year all-cause mortality better than models using clinical metrics alone^11^. Similarly, Mohammad and colleagues developed and validated a neural network to predict 1-year mortality and re-admission for heart failure after incident myocardial infarction with greater discrimination than the GRACE2.0 score^12^. However, the majority of the secondary risk prediction models based on ML have not used time-to-event analysis. One notable exception is the model presented by Steele et al, which however have not been externally validated^10^. By not using survival analysis, previous ML-based analyses omit data points with incomplete follow-up (censoring) and thereby effectively prevents a model from distinguishing between “died after a week” and “died after 10 months” which we believe is of obvious clinical interest.

To overcome these limitations, we describe the development and validation of a neural network-based survival model, PMHnet, for predicting all-cause mortality in patients with IHD using 584 different features extracted from population-wide healthcare registries and complete EHRs. We identified several influential features which previously have been omitted from risk prediction of patients with IHD.

## Methods

### Data foundation

The algorithm PMHnet was developed using a cohort constructed from the Danish National Patient Registry (NPR) and the Eastern Danish Heart Registry (EDHR)^14^. The EDHR contains structured information on all coronary artery angiographies (CAGs) performed in the Capital Region of Denmark and Region Zealand. The cohort was linked to a population-wide EHR database that covers Eastern Denmark from 1^st^ of January 2006 to the 7^th^ of July 2016 (BTH), and genotype data from the Copenhagen Hospital Biobank Cardiovascular Diseases study (CHB-CVD)^15–17^. The BTH dataset fully covered Eastern Denmark (2.6 million patients). Outcomes were obtained from the Central Person Registry and the Danish Register for Causes of Death^18, 19^. Data sources were linked using encrypted Danish personal identification numbers^19^.

### Selection criteria and model development

First, we identified all adult Danish citizens (>18 years of age) in NPR with an ICD-10 code for IHD (I20-I25) who had undergone their first CAG between Jan 1, 2006, and Jun 1, 2016, demonstrating one-, two-, or three-vessel disease (1–3VD) or diffuse atheromatosis (DIF). Vascular disease is here defined as stenosis above 50%^20^. For patients fulfilling these criteria (n=39,746), we used the date of the CAG as the index date and included five years of follow-up. Patients were followed until either death or censoring, whichever came first. Using the hold-out method, the derivation data was randomly divided into a training set (n=34,746) and a test set (n=5,000) used for model development and independent assessment of performance, respectively^21^ (Figure 1, Table 1).

**Figure 1:**
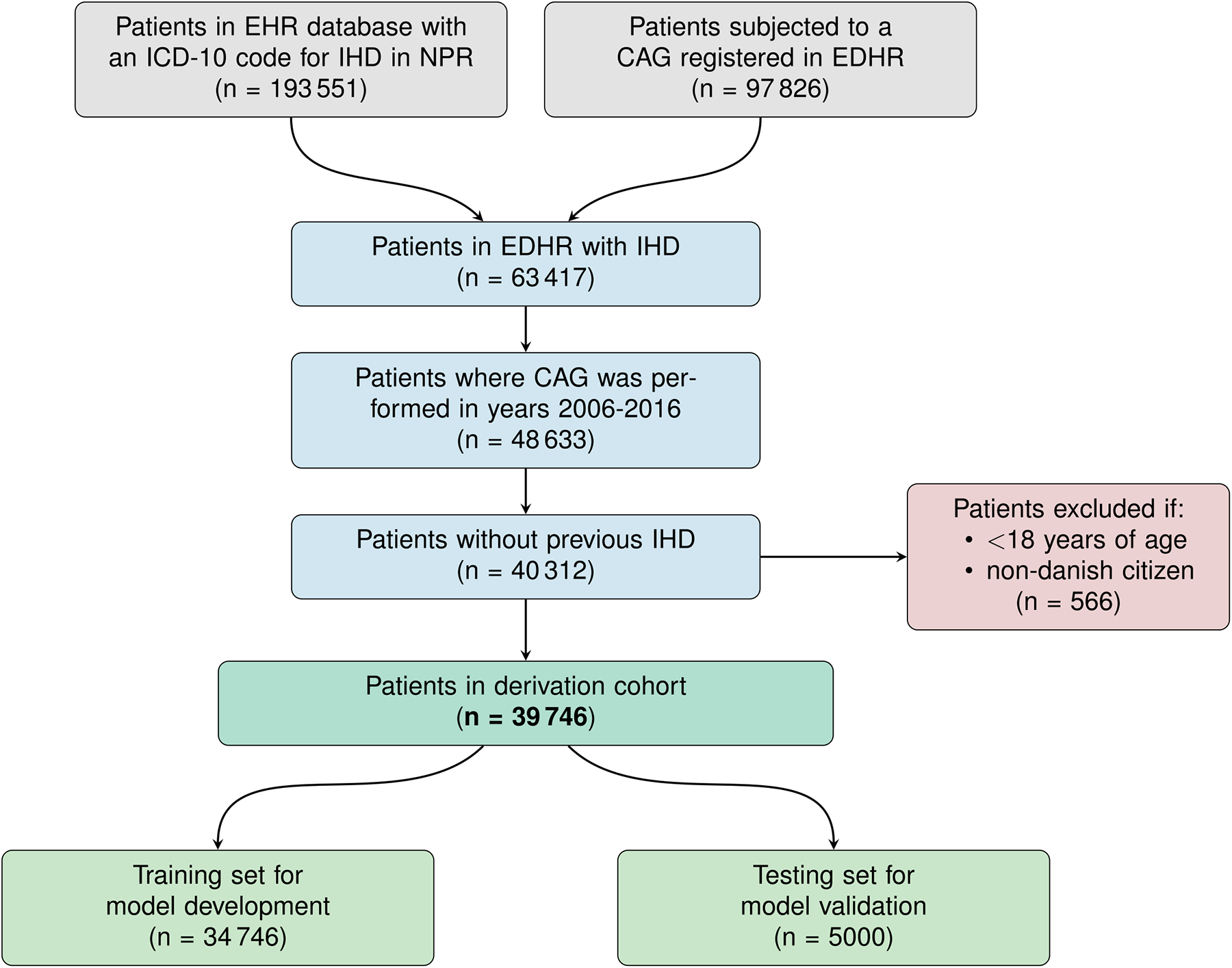
Flowchart of inclusion of patient with ischemic heart disease in the training and test set for the PMHnet. Flowchart showing how the derivation cohort was identified based on data from NPR and EDHR. CAG: Coronary arteriography. EHRs: Electronic health records. EDHR: Eastern Danish Heart Registry. ICD-10: International classification of diseases, 10th revision. IHD: Ischemic heart disease. NPR: Danish National patient registry.

**Table 1:**
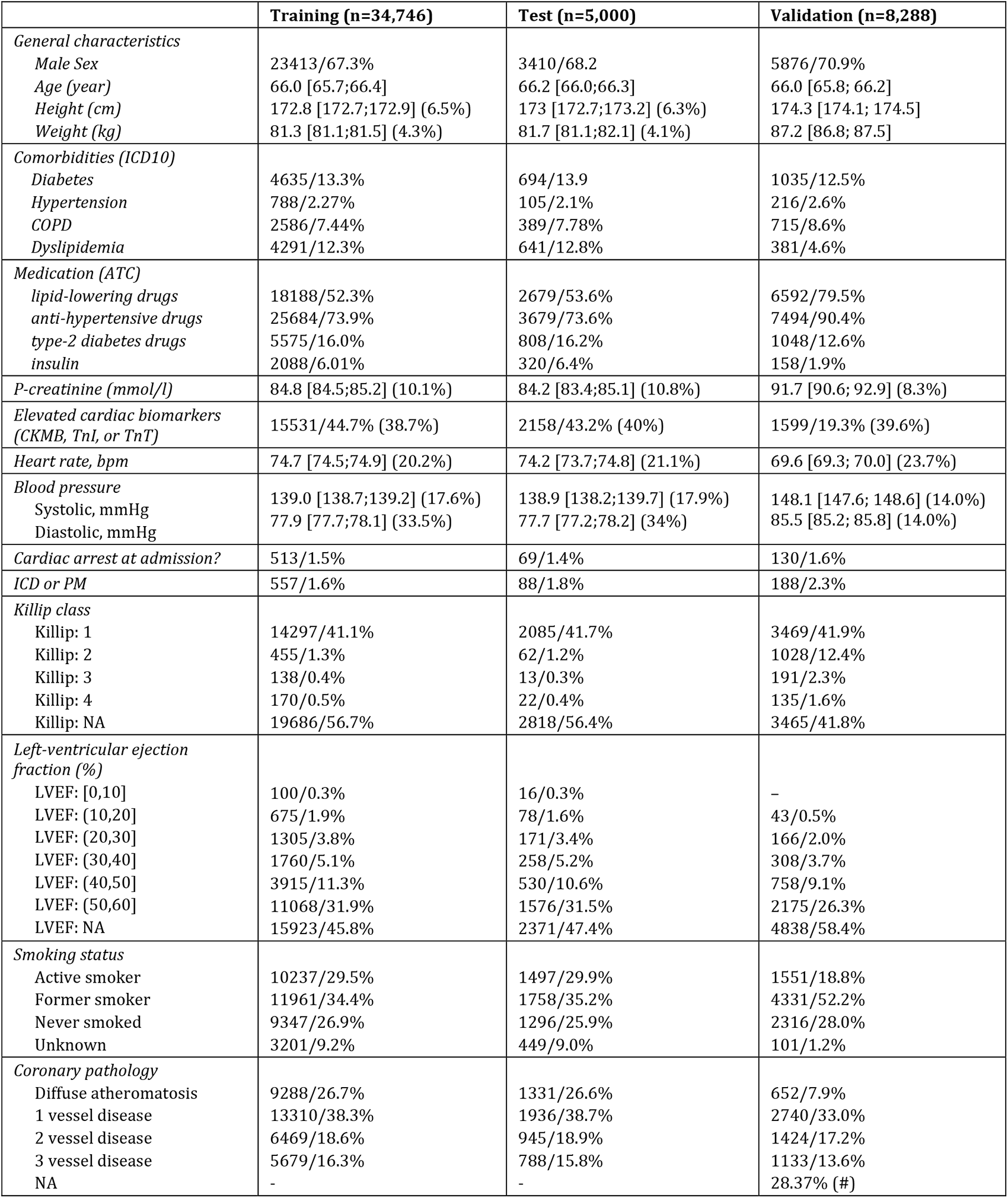
Cohort characteristics for training, test, and external validation set. For continuous features (age, height, etc.) the mean is given along with 95% bootstrap confidence intervals (CI) and, if applicable, the amount of missingness in parentheses – mean [low, high] (missingness). The mean and CI are calculated using only the non-missing features. For categorical features the raw counts and relative frequencies are both specified. The comorbidities are defined from the ICD-10 codes that had been assigned to a given patient prior to the index date. Medication is defined from prescriptions prior to index date. Lipid-lowering drugs: C10, anti-hypertensive drugs: C02, C03, C07, C08 and C09, type-2 diabetes drugs: A10B, insulin: A10A. See supplementary methods for further details. (#): Diffuse atheromatosis could not be defined with complete certainty for 28.4% of the Icelandic data, and coronary pathology was therefore set as NA in such cases. COPD: Chronic obstructive pulmonary disease, ICD: Implantable cardioverter-defibrillator, LVEF: Left-ventricular ejection fraction. PM: Permanent pacemaker.

For each of the 39,746 patients, we reduced the available features prior to index event (i.e. first CAG between Jan 1, 2006, and Jun 1, 2016) to a smaller set based on prevalence such that e.g., a diagnosis code could be found in at least 5% of the training set. The final set of 584 features was separated into five different categories: *ClinicalOne* (8 features), *ClinicalTwo* (15 features)*, Diagnoses (322 features)*, *Procedures (154 features)*, and *Biochemical* (85 features) (Table 2). *ClinicalOne* included the same eight input features as used by GRACE2.0 and *ClinicalTwo* had 14 additional clinical features (Table 2) that were selected based on availability. Features were defined using data recorded prior to the index date, except for creatinine, cardiac biomarkers for ischemic heart disease, and blood pressure where high missingness led us to allow measurements obtained after the CAG in cases of missingness (7-day threshold for cardiac biomarkers, and 31-day threshold for the others). *Diagnoses* included ICD-10 codes registered in NPR and similarly, *Procedures* consisted of procedure codes (surgery and examinations such as X-rays) registered in NPR. *Biochemical* contained results of in-hospital blood tests. Additional details on feature extraction, missingness, and pre-processing can be found in supplementary methods. The amounts of missingness across the training and test set have been tabulated in Table S1. Missing values were left missing and encoded as such in PMHnet.

**Table 2:** Input features used for model development. The different features were organized in five different categories each representing different domains. The clinical characteristics were divided into two subgroups where ClinicalOne contains the features used in the GRACE2.0 score.

| Category | Features |
| --- | --- |
| *ClinicalOne*<br>(CRACE2.0) | Age, pulse, systolic blood pressure, cardiac arrest at presentation (yes/no), abnormal cardiac enzymes (yes/no), Killip-class, creatinine, ST-segment deviation (yes/no) |
| *ClinicalTwo* | Abnormal ECG (yes/no), CCS class, diastolic blood pressure, coronary artery dominance (R/L/B), familial IHD (yes/no), height, weight, ICD-device or PM (yes/no), ischemia test, LVEF, NYHA class, sex, smoking status, coronary pathology, |
| *Diagnoses* | 322 different level-3 ICD-10 diagnosis codes. |
| *Procedures* | 154 different NOMESCO procedure codes corresponding to various radiological examinations and surgical procedures |
| *Biochemical* | 85 different lab tests with results categorized as *below*, *within*, or *above* the reference range |

### External validation data from Iceland

For the external validation cohort, we identified Icelandic adults who had undergone CAG at the only interventional cardiology center in Iceland, Landspitali– The National University Hospital in Reykjavík^22^. We obtained data collected prospectively between January 1, 2007, and December 31, 2017. Information on ICD-10 diagnoses and procedure codes were aggregated from the Landspitali, from registers kept by the Directorate of Health: The Register of Primary Health Contacts, the Register of Contacts with Medical Specialists in Private Practice and the Causes of Death Register, as well as at recruitment for deCODE studies. Biochemical assay measurements were obtained from the three largest clinical laboratories in Iceland, with measurements performed at: (i) Landspitali; (ii) The Laboratory in Mjodd, Reykjavik, Iceland; and (iii) Akureyri Hospital, the regional hospital in North Iceland.

### Polygenic risk scores

Polygenic risk scores (PRSs) for patients with genotypes available through the CHB-CVD^17, 23^ (31.4% of the cohort) were calculated using the LDpred2 framework, implemented in the R package bigsnpr (v1.5.2) with R version 3.5.052^24^. PRSs were calculated based on GWAS summary statistics data from 19 traits relevant for cardiometabolic health, obtained from 17 GWAS meta-analyses. List of meta-analyses and details on the PRS calculations is included in the Supplementary Material.

### Machine learning model architecture and development

To model time-to-event data and allow for censoring, we used the generic discrete-time survival model for neural networks described by Gensheimer and Narasimhan^25^. In this model, follow-up time is divided into a fixed number of intervals and the model estimates a conditional hazard for each interval, i.e., the probability of dying in that time interval given that the patient is still alive at the end of the preceding interval. PMHnet uses 30 intervals separated in time such that event times in the training data are evenly distributed across all intervals. To obtain predictions between breakpoints in the discretization grid, we assumed that the probability density function was constant in each time interval, and we thus interpolated using a piecewise linear function^26^. The implementation applied the PyTorch machine-learning framework using the authors’ Keras version as a reference^25^.

We used a feed-forward neural network and tested various hyperparameters. The output layer was a fully connected sigmoid activated layer that outputs conditional hazards for each of the 30 different time points. We added dropout to each of the hidden layers to regularize the network and prevent over-fitting. The number of layers, neurons, learning-rate, and dropout rate for each layer were fine-tuned through hyperparameter optimization using the Optuna optimization framework, with a five-fold cross validation^27^. The hyperparameter search space and the best trial for the complete model is included in table S3. The neural networks were trained using stochastic gradient descent, with a constant learning rate, to minimize the negative log-likelihood.

### Model evaluation and validation

Using the hold-out test set and the external validation data from Iceland, performance of PMHnet was evaluated through assessment of both model discrimination and calibration. We used time-dependent area under the receiver operating characteristic curve (tdAUC) as the main measure of discrimination, but also calculated the Brier score that can be used to assess both discrimination and calibration^28–30^. Calibration was also analyzed graphically by comparing the predicted risks with the estimated actual risks^28^. The Score function from the riskRegression R package was used to compute performance measures and compare models. For comparisons between two competing models, the Score function gives p-values that correspond to Wald tests on the standard errors obtained using an estimate of the influence functions following Blanche et al. ^30^.

To benchmark PMHnet we calculated the GRACE2.0 for all patients in the hold-out test set, using the GRACE2.0 webtool. We extracted the javascript source code for the GRACE2.0 webtool using the developer tools in Google Chrome. The javascript code was then manually converted to an R package for automatized computation of GRACE2.0 on the entire cohort. The eight variables used in GRACE2.0 were available for 51.4% of the cohort. Since GRACE2.0 does not allow for missing features, we imputed missing variables using the missForest R package^31^. Imputed values were only used for calculating the GRACE2.0 score. In addition to the conventional GRACE2.0 score, we re-fitted the GRACE2.0 score to our training data using the PMHnet architecture. This corresponds to model_2 in Figure 4 that only uses the features from *ClinicalOne* as its input.

For independent validation of PMHnet we used, as described above, Icelandic EHR data from 8,287 patients. Of the 584 features identified in the Danish derivation cohort, we found matching data for 404 features in the Icelandic data. A down-scaled model was re-trained on the Danish training set to make comparisons.

### Explainability and effect of missing features

To investigate the impact of different features on model predictions and to provide model explanations, we calculated Shapley additive explanation (SHAP) values for all features and patients in the training set using the SHAP python package^32^.

To assess how resilient the model was in the event of missing data, missingness was introduced in the test data by replacing all values of a given feature with the median value of that feature in the training data. The predictions were then compared with the predictions of PMHnet. Resiliency was then quantified using change in tdAUC (discrimination) and Brier scores (calibration), where the predictions of PMHnet were compared to that of model with artificially introduced values (a total of 584 comparisons).

### Statistical analysis

Categorical features are reported as counts (%) and continuous features as mean [95% CI], 95% CIs are obtained from standard deviations or through bootstrapping. Time-dependent AUCs and Brier scores were calculated using the riskRegression R-package^28^. Likewise, model comparisons were obtained from the same package. All statistical analyses and visualizations were performed using R version 4.1.

### Data access and ethics approvals

The study was approved by The National Ethics Committee (1708829, ‘Genetics of CVD’—a genome-wide association study on repository samples from CHB), The Danish Data Protection Agency (ref: 514-0255/18-3000, 514-0254/18-3000, SUND-2016-50), The Danish Health Data Authority (ref: FSEID-00003724 and FSEID-00003092), and The Danish Patient Safety Authority (3-3013-1731/1/). Danish personal identifiers were pseudonymised prior to any analysis.

The study was approved by the Data Protection Authority of Iceland and the National Bioethics Committee of Iceland (VSN-15-114). Icelandic participants that donated biological samples provided informed consent. Personal identities of the participants were encrypted with a third-party system provided by the Data Protection Authority of Iceland.

Study design, methods, and results were reported in agreement with the TRIPOD statement^33, 34^ and following the STROBE recommendations^35^.

## Results

The derivation cohort of 39,746 Danish patients with IHD were randomly subdivided into a training (N=34,746) and a test set (N=5,000) (Table 1). At inclusion the patients mean age (95%-CIs) was 66.0 years [65.7; 66.4] (67.3% males) in the training set and 66.2 years [66.0;66.3] (68.2% males) in the test set. The distribution of the degree of coronary artery disease was similar in the two groups (distributions of patients presenting with one-, two-, or three-vessel disease or diffuse atherosclerosis, respectively).

The Kaplan-Meier estimate of five-year survival (all-cause) was 81.8% [81.4; 82.2] for the training set and 82.5 [81.3; 83.6] for the test set (Figure S1, Table S2). The restricted mean follow-up time was 1,635 days (±2.58) for the training set and 1,635 days (±6.49) for the test set.

### PMHnet model predictions

In the internal validation using the hold-out test set, the complete PMHnet model had tdAUCs of 0.88 [0.86; 0.90] at six months, 0.88 [0.86; 0.90] at one year, 0.84 [0.82; 0.86] at three years (Figure 2), and 0.82 [0.80; 0.84] at five years. In comparison, the corresponding values for the conventional GRACE2.0 score on the same dataset were 0.77 [0.74; 0.80] at six months, 0.77 [0.74; 0.80] at one year, and 0.73 [0.71; 0.75] at three years. For the re-fitted GRACE2.0 score, tdAUCs were 0.79 [0.76; 0.83] at six months, 0.78 [0.75; 0.81] at one year, and 0.76 [0.74; 0.78] at three years. Since GRACE2.0 features had to be imputed for 48.6% of the population, we also evaluated the performance on the subset without missingness, which were largely the same (Figure 3, dashed line). GRACE2.0 is not designed for providing predictions after three years and was therefore not evaluated beyond that time-point. The difference in tdAUCs between PMHnet and either of the two GRACE2.0 models was significant at each of the three prediction horizons (Table 3).

**Figure 2:**
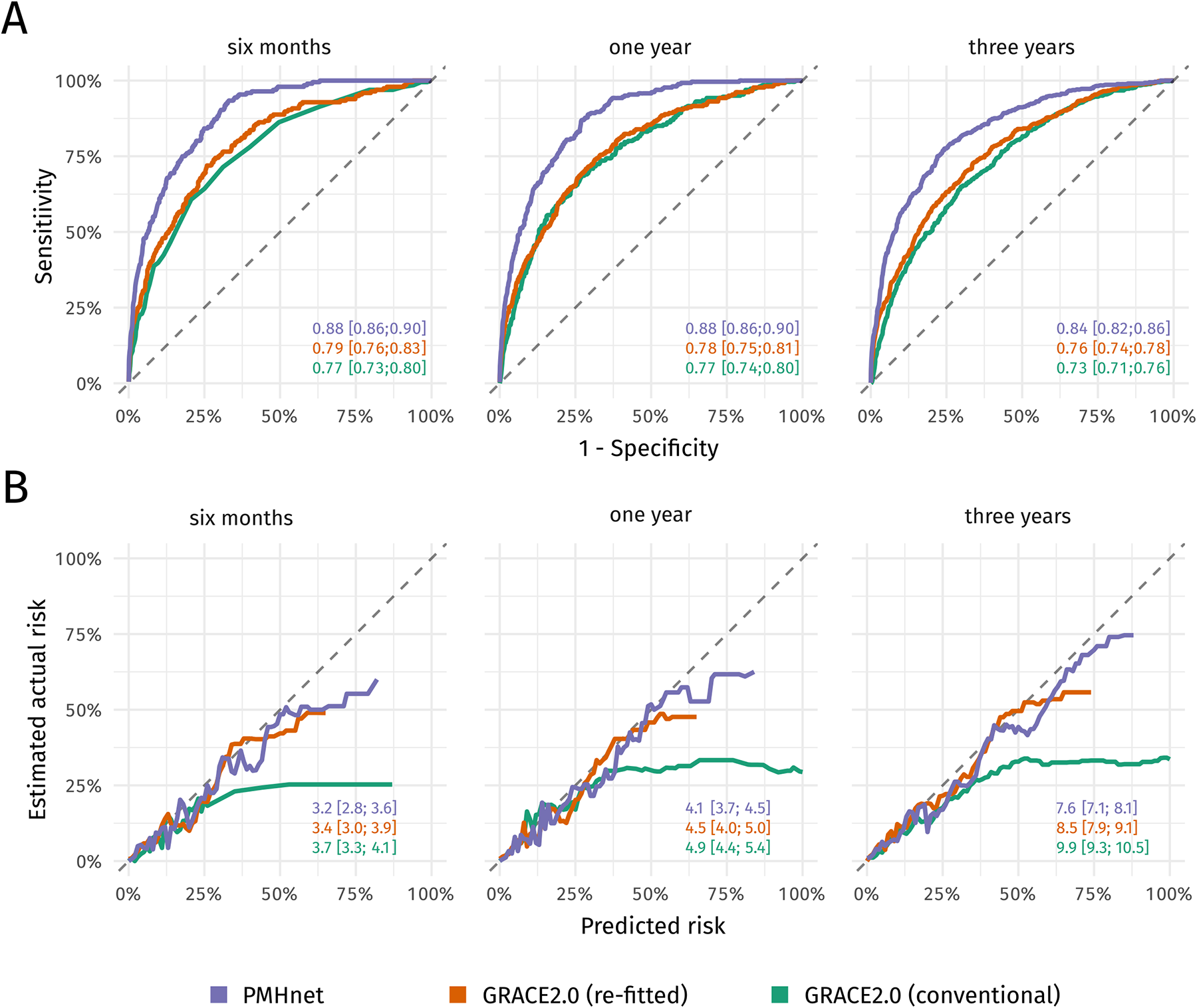
Model performance of PMHnet and the GRACE2.0 score. A) Time-dependent receiver operating characteristics (ROC) curves at three different prediction horizons for PMHnet, GRACE2.0 (re-fitted), and GRACE2.0 (conventional). Labels show the time-dependent area under the ROC curves (AUC). GRACE2.0 (re-fitted) is a model that uses the GRACE2.0 input features but uses the PMHnet architecture and is trained using our training data. B) Calibration curves showing the relation between predicted risk and the estimated actual risk. Labels show the Brier score for each of the three models. Lower scores are associated with better calibration and discrimination of predictions.

**Figure 3:**
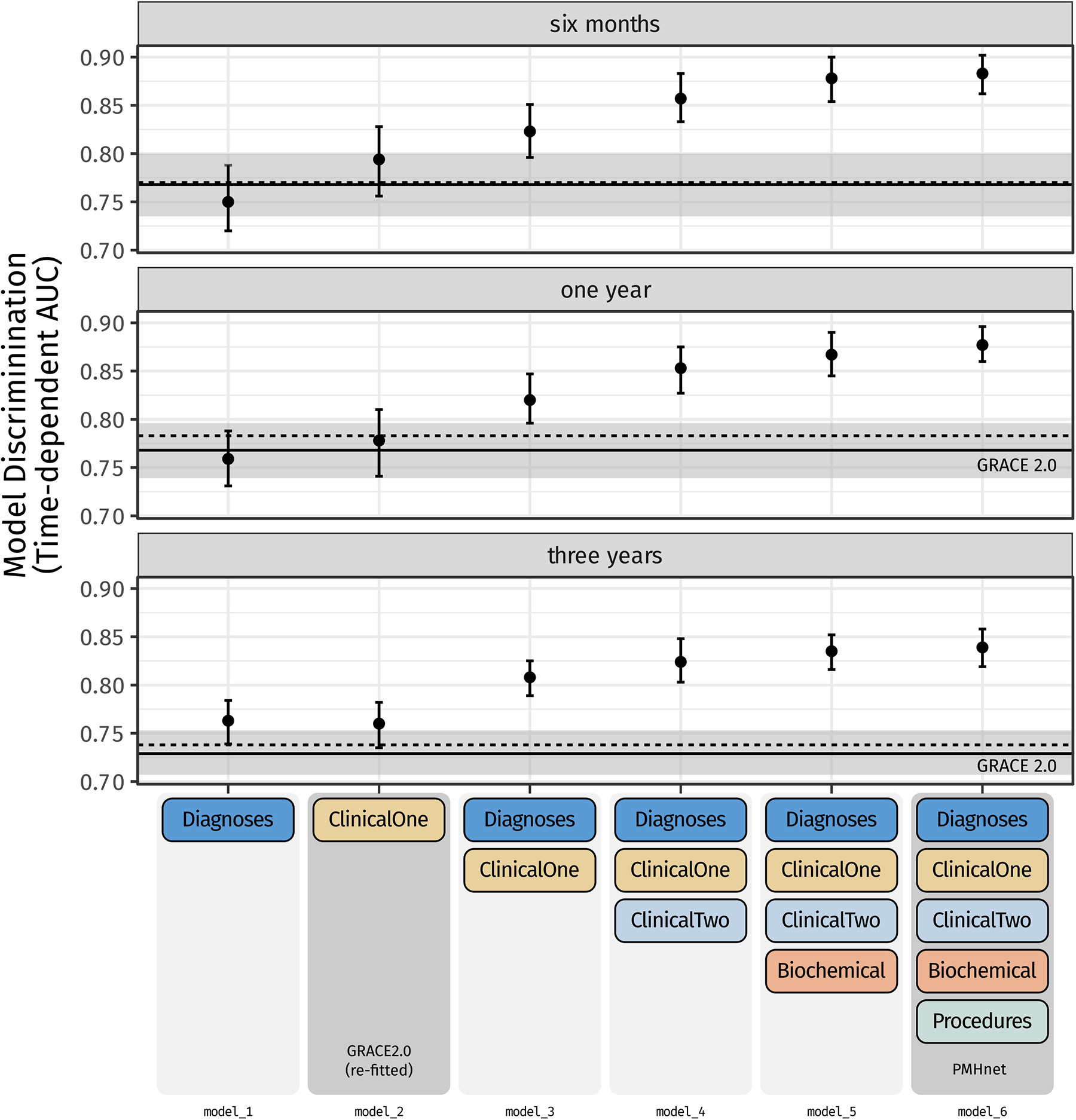
Model discrimination with increasing number of feature categories. Time-dependent area under the curve (tdAUC) for various intermediate PMHnet models (and the final one (model 6)) at six months, one year, and three years after the index coronary angiography. Discrimination was evaluated using the hold-out test set. The colored boxes represent the different feature categories that were used as model input in the different models. Horizontal reference lines show the model discrimination of the GRACE2.0 score on the same data. The solid line is the tdAUC of GRACE2.0 on all patients and the dotted line is the tdAUC on the subset of patients where none of the GRACE2.0 input features were missing.

**Table 3:**
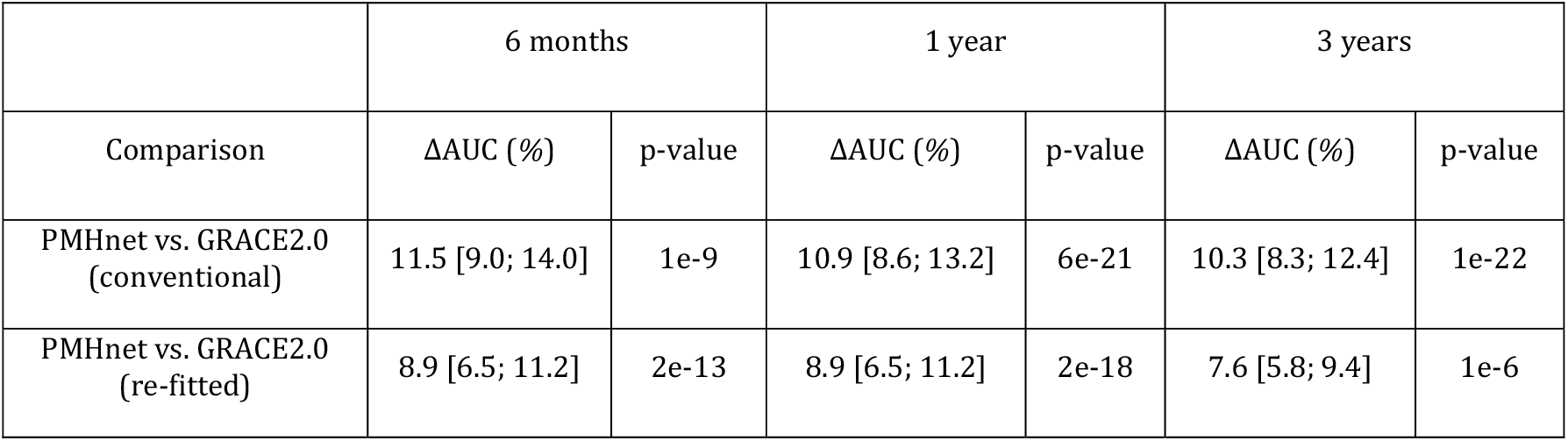
Difference in discrimination between PMHnet and the GRACE2.0 score. For ΔAUC, we obtain 95% CIs (in brackets) and p-values from the Score function in the R package riskRegression^38^.

|  | 6 months |  | 1 year |  | 3 years |  |
| --- | --- | --- | --- | --- | --- | --- |
| Comparison | ΔAUC (%) | p-value | ΔAUC (%) | p-value | ΔAUC (%) | p-value |
| PMHnet vs. GRACE2.0 (conventional) | 11.5 [9.0; 14.0] | 1e-9 | 10.9 [8.6; 13.2] | 6e-21 | 10.3 [8.3; 12.4] | 1e-22 |
| PMHnet vs. GRACE2.0 (re-fitted) | 8.9 [6.5; 11.2] | 2e-13 | 8.9 [6.5; 11.2] | 2e-18 | 7.6 [5.8; 9.4] | 1e-6 |

As an additional visual test of discrimination, we constructed five different risk strata using the 5-year predicted survival (defined as 90%, 75%, 50%, and 25%) and examined the observed survival (Figure 4), which showed good separation between the five strata. PMHnet was found to be well-calibrated as seen from Figure 2B and the calculated Brier scores of 3.2% [2.8; 3.6] at six months, 4.1% [3.7; 4.5] at one year, 7.6% [7.1; 8.1] at three years, and 11.3% [10.5; 12.0] at five years.

**Figure 4:**
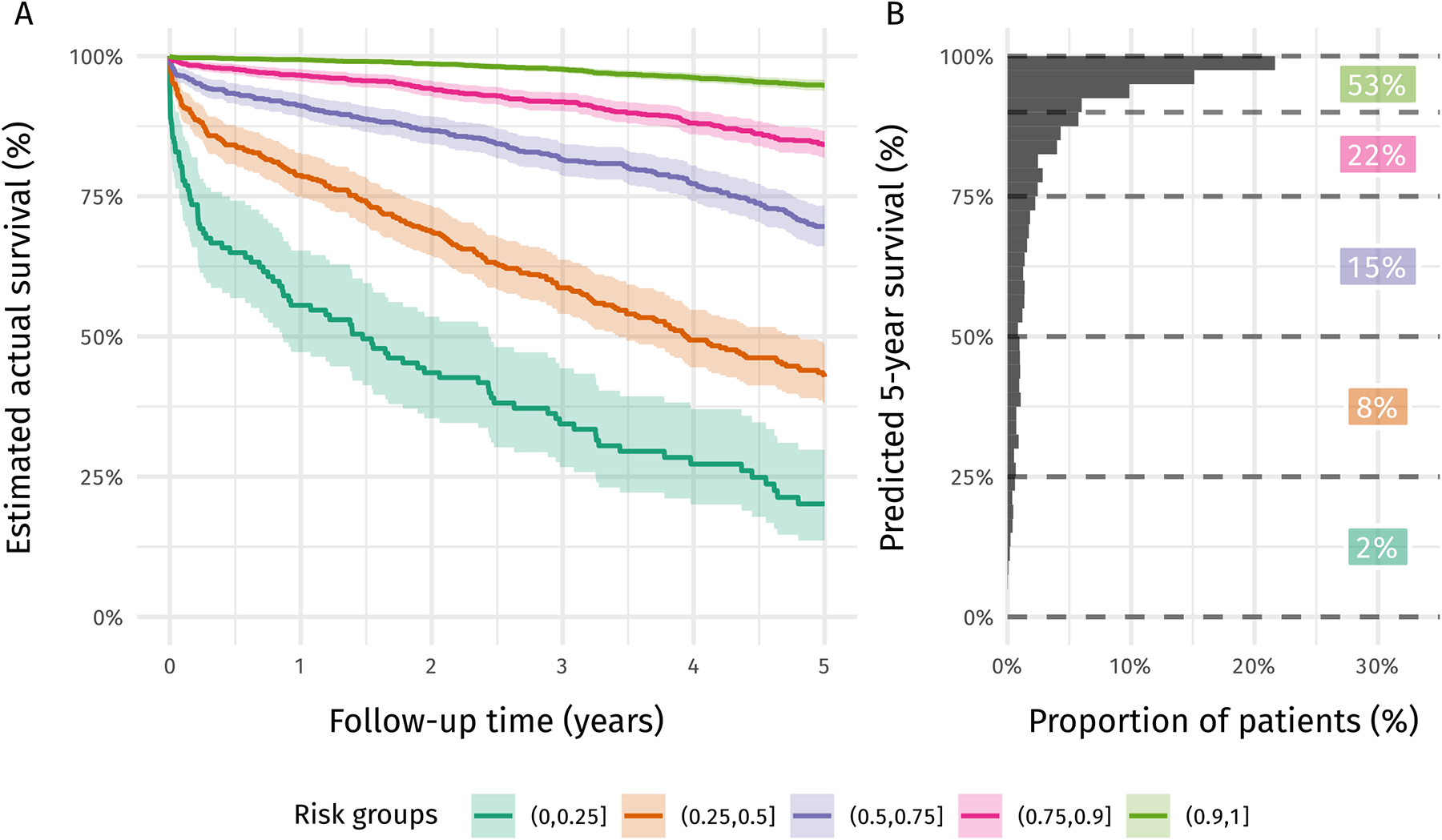
Observed survival across 5-year predicted risk groups. A) Estimated actual survival (Kaplan-Meier estimates) for patients in the test set manually stratified into five different risk-groups depending on the predicted survival at five-years by PMHnet. B) Distribution of PMHnet 5-year predicted risk. Vertical lines show the cut-offs that are used to define the risk strata used in A).

In comparison, GRACE2.0 had Brier scores of 3.7% [3.3; 4.1] at six months, 4.9% [4.4; 5.4] at one year, 9.9% [9.3; 10.5] at three years. Re-fitting GRACE2.0 with the PMHnet architecture considerably improved the calibration as evident from the calibration curve which was comparable to that of PMHnet (Figure 2B). The differences in three-year predicted risk between PMHnet and the two GRACE2.0 scores for all patients in the test set are shown in Figure S2.

### External validation of PMHnet

For external validation the cohort of 8,287 patients from Iceland was used (Table 1, *validation set*). Due to data availability a modified, and down-scaled version of PMHnet was used (404 features) on the Icelandic data. The tdAUCs were 0.86 [0.84;0.90] at six months, 0.84 [0.81;0.87] at one year, and 0.81 [0.79;0.83] at three years. In comparison, the performance of the down-scaled version on the Danish (internal, hold-out) test set was 0.87 [0.85;0.90] at six months, 0.87 [0.85;0.89] at one year, and 0.82 [0.80;0.85] at three years. The predictive performances were highly concordant, while model calibration was slightly worse in the Icelandic data (Figure S3).

Influence of the different input feature categories on the PMHnet prediction

To assess the importance of the different input feature categories systematically, we trained five intermediate survival models using the PMHnet architecture (Figure 3). For example, *model-1* used diagnosis codes as input features only, *model-2* used the clinical variables known from GRACE only. The other intermediate versions were trained using different combinations of the feature categories. Figure 3 shows the model discrimination at the three different prediction horizons for the six different models fitted using the PMHnet architecture. The performance of the intermediate model based on diagnosis codes only (*model-1*) was similar to the performance of the re-fitted GRACE2.0 model (*model-2*). Interestingly, combining diagnosis codes with the GRACE2.0 clinical features (*Diagnoses* + *ClinicalOne*) in *model-3*, there was an overall increase in AUC at three years, which would suggest a synergistic effect. With the addition of the *ClinicalTwo*-data, the ΔAUC between *model-3* and *model-4* was 3.4 [1.8; 5.0] at six months, 3.2 [1.8; 4.7] at one year, and 1.3 [0.3; 2.3] at three years. Similarly, adding *Biochemical* to the input features in *model-5* associated with significant ΔAUCs of 2.1 [0.2; 3.6] at six months, 1.4 [0.2; 2.7] at one year, and a non-significant ΔAUC of 0.9 [-0.1; 1.9] at three years. The gain in discrimination from *model-5* to the complete *model-6* (PMHnet) by adding *Procedures* to the input features was only significant at the one-year prediction horizon.

### Testing inclusion of polygenic risk scores in PMHnet

For 37.4% of the cohort, we had genotype information available and used that to calculate 19 different polygenic risk scores (PRS) that all related to different cardiometabolic traits. Limiting both the training set and the test set to only individuals with genotype data (37.4%), we tested adding the PRS scores to *model-1* (*Diagnoses*) and *model-2* (*Diagnoses* + *ClinicalOne*) and evaluated the model performance (Figure S4). Addition of the PRS did not significantly improve either model discrimination at any time-point (Figure S4A).

### Explainability analysis using SHAP

We performed SHAP-analyses of the five-year model predictions to quantify the impact of the different features on PMHnet predictions. SHAP is a technique rooted in cooperative game theory that provides an estimate of feature impact on the model output. It quantifies the contribution of each feature to the prediction outcome, allowing for a better understanding of feature importance and model behavior. By considering the interactions and dependencies between features, SHAP analysis provides insights into the specific factors influencing the model’s decision-making process and aids in identifying key drivers and relationships within the model^36^. Across the five different feature categories, biochemical test results and diagnosis codes were most impactful, while the category *ClinicalOne* was least impactful (Figure 5A). The most impactful diagnosis code was chronic obstructive pulmonary disease (ICD10: J44) and thus ranked higher than classical risk factors for IHD, such as type 2 diabetes. At the feature level, age, the number of affected vessels (1–3VD, or DIF), and smoking were on-average the most predictive features. The top-25 features in terms of average model impact (average magnitude of SHAP-value), included ten features from *Biochemical*, nine from *ClinicalTwo*, three from *ClinicalOne*, two from *Diagnoses*, and one from *Procedures* (Figure 5A). To further examine the impact on model prediction for the two most impactful features, number of affected *vessels* and *age*, we constructed SHAP dependence plots (Figure 5B)^37^. For *age* we observed non surprisingly that higher age pulled the prediction towards non-survival, and that age was estimated to add/remove anywhere between -25 and 20 percent points to the predicted 5-year survival. Similarly for *vessels*, more affected vessels impacted the predicted survival negatively.

**Figure 5:**
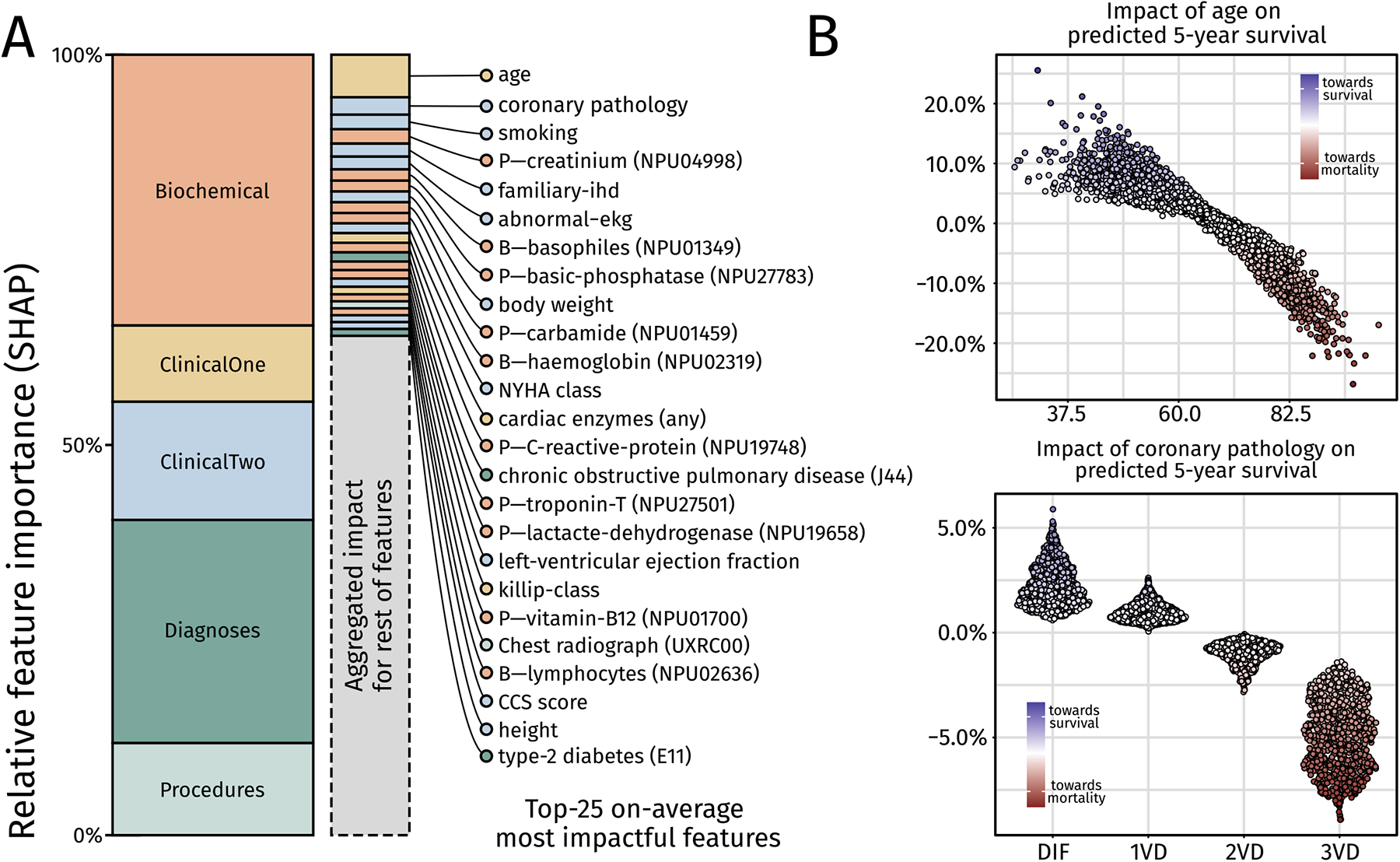
Overview of feature importance. Summary of the results from SHAP analysis on the model predictions at five years on patients in the Danish test set. A) Left: Relative feature importance aggregated across the five different feature categories. Biochemical test results and diagnoses were found to affect the model prediction the most. Right: Relative feature importance for all singular features included in the model. Features arranged according to SHAP-values and labels are included for the top 25-most impactful features. Color of features correspond to the feature category in which they belong. SHAP: SHapley Additive exPlanations. B) Relationship between age and SHAP-value with each point showing the SHAP-value for age for a patient in the test-set, and relationship between coronary pathology (e.g. vessel status) and impact on model prediction.

For both features, we noted that identical feature values not always impacted the survival to the same extent. This vertical dispersion in both plots represent interaction effects with the other included features^37^. Although our SHAP analyses reveal that many features have lower impact relative to the most impactful features, the aggregated sum of the many low-impact features (560 lowest) outweighs many of the well-known risk-factors (25 highest). For this reason, we did not attempt to limit the number of included features. Extending the analysis of feature-level model impact to the rest of the top-25 features, we constructed a summary plot of the SHAP-values that shows the distribution of model impacts across feature values (Figure 6).

**Figure 6:**
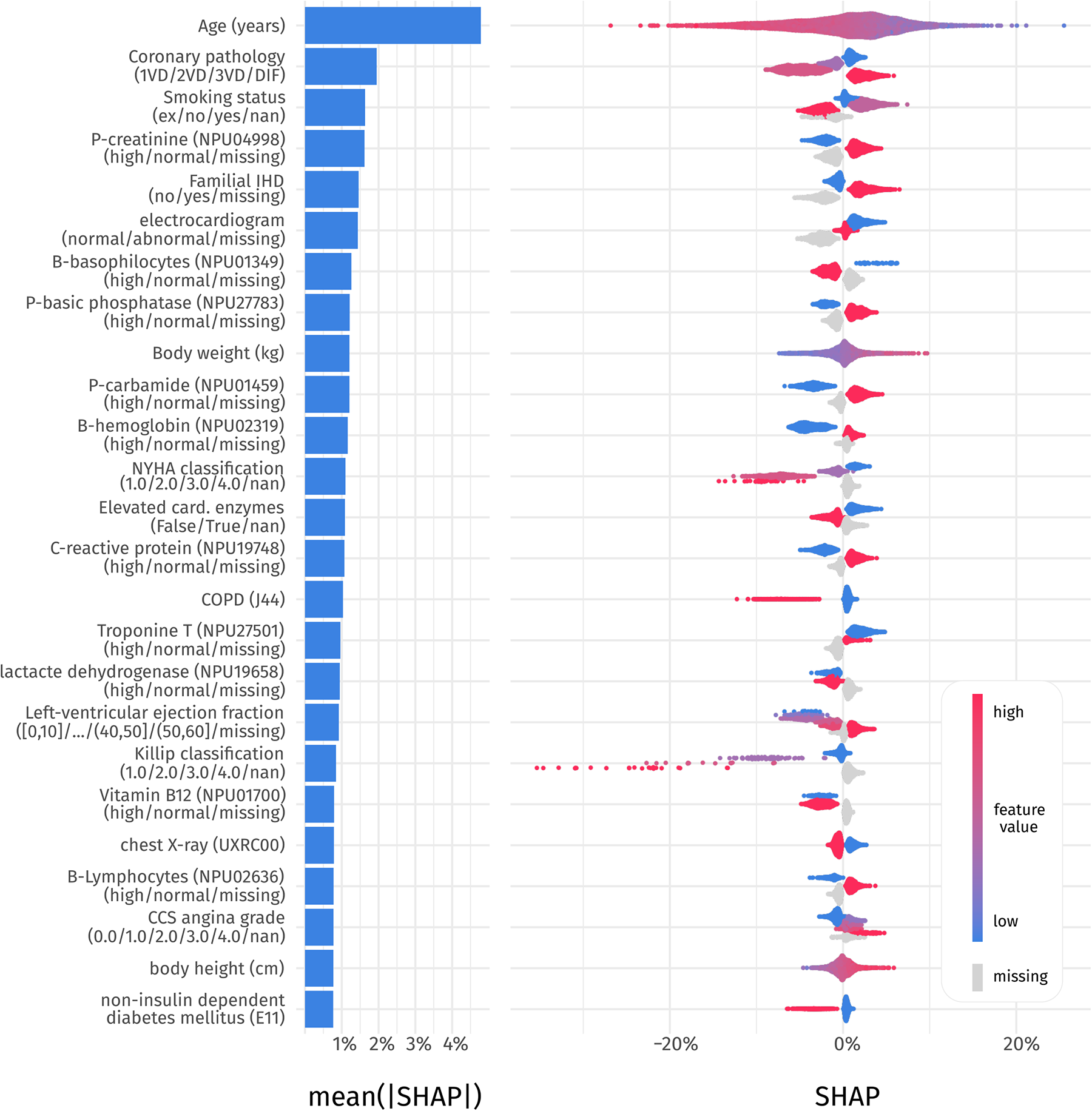
SHAP summary plot for top 25 most impactful features. Left: average magnitude of model impact. Y-axis labels specifies the feature name and inside parentheses is shown the unit or the factor levels, for continuous and categorial features, respectively. Right: Distribution of feature impacts across the test set. For continuous features, the colors correspond to the feature value ranging from blue (smallest) to red (largest). For categorical features, the factor levels are colored from blue to red. Grey indicates missingness.

### Patient-level feature importance

Finally, we also generated individual explanations for three example patients in the test set and show the SHAP values for the nine most impactful features (Figure 7). The patients were randomly selected from the subsets of patients with a prediction in the intervals (0.25; 0.5], (0.5, 0.75], and (0.75; 1]. For *patient 1*, with the worst 5-year prognosis, *age* was not among the nine most impactful features. Instead, the explainability algorithm highlights diagnosis codes and biochemical values. For *patient 2* with a 5-year predicted risk of 73%, the most impactful feature was age (78 years), which pulled the prediction towards mortality. The impact of age was however largely cancelled by *rest*, which constitutes the aggregated sum of all the features not among the nine most predictive. For *patient 3*, the feature *age* was again highlighted as the most impactful, but in this case, it impacted the prognosis positively. Apart for a history of cigarette smoking, none of the highlighted features had negative impact on the prognosis.

**Figure 7:**
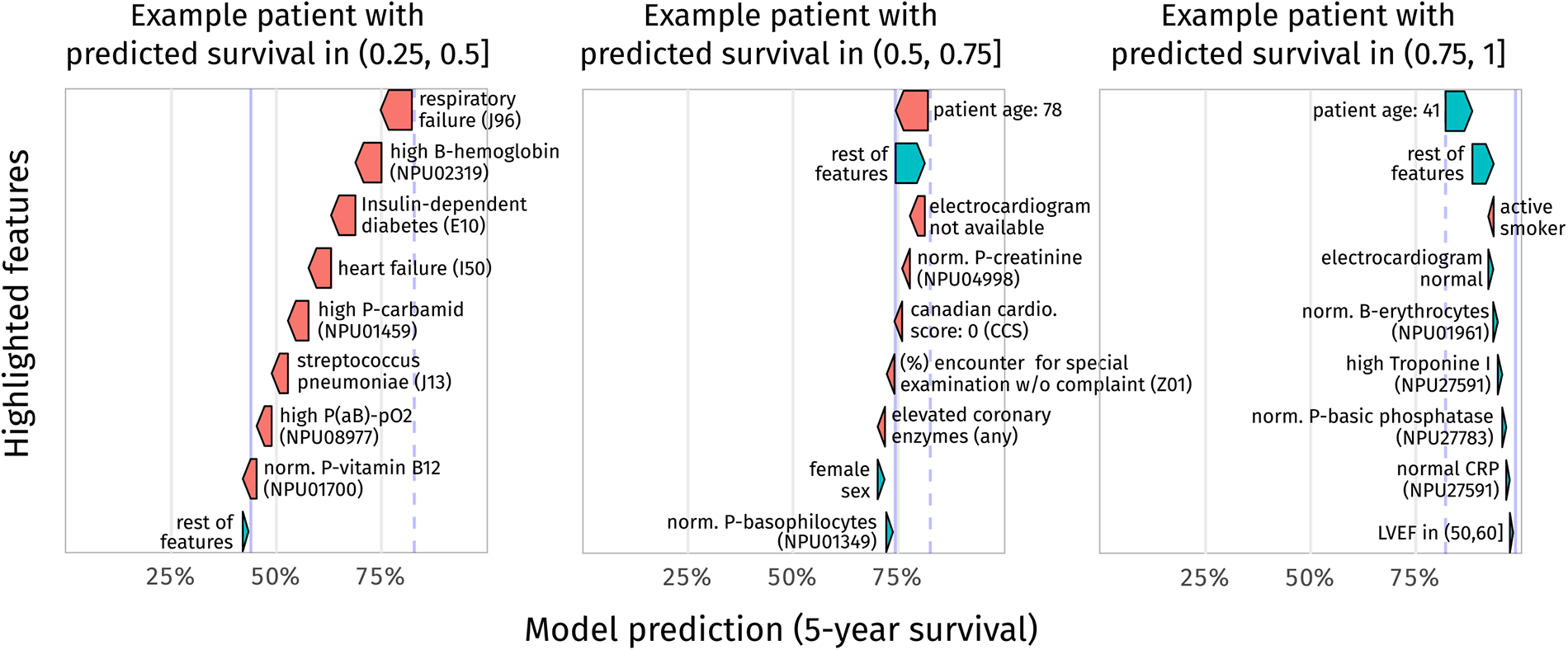
Patient-level model explanations with SHAP. Model predictions for three different representative patients with a predicted 5-year survival of 42%, 73%, and 98% with SHAP-explanations showing the estimated impact on model prediction. The patient data have been slightly adjusted to make them non-identifiable. Blue represents features that contribute positively to the prediction. Red represents features that contribute negatively to the prediction. SHAP: SHapley Additive exPlanations.

## Discussion

In this study, we developed a feature-rich neural network-based survival algorithm, PMHnet, for prediction of all-cause mortality in patients with IHD using data from 34,746 Danish patients. With the aim of providing predictions that can be used to guide treatment and care, the model was developed to operate with an index date immediately after the diagnosis-confirming coronary angiography and with a prediction horizon of five-years. The model was tested using data from 5,000 Danish patients and externally validated using data from 8,288 Icelandic patients. We found that PMHnet had excellent discrimination with tdAUCs ranging from 0.88 at six months to 0.82 at five years. Similar results were found on the external Icelandic data, which confirms that the model and its deep feature foundation generalized well to novel patients and a different healthcare setting. Evaluated on the Danish data, we found the model to be well-calibrated with predicted probabilities accurately reflecting the observed proportions, also in different risk strata.

To aid the clinical interpretation of model predictions, we used SHAP-values to highlight the most impactful features and to explain how the different features affect the prediction for the individual patient. Model explainability is important for evaluating the model output and is paramount for the clinical adaption of any ML-model^38^, including focus on features that are clinically actionable, i.e. modifiable versus non-modifiable factors.

Compared to the GRACE2.0 score, which is widely considered the gold-standard risk-stratification tool in current clinical use for predicting mortality after acute coronary syndrome (ACS)^39^, PMHnet had superior discrimination and calibration. However, it is important to note that there are differences between the intended patient populations for the two models. The GRACE2.0 score has been developed using a derivation cohort of patients with STEMI, n-STEMI, and unstable angina with time of initial admission as time-zero^5^. In contrast, our study used time at coronary angiography as its baseline and was applied to all patients with IHD and coronary artery pathology ranging from diffuse atheromatosis to three-vessel disease. The validity of GRACE for a cohort with such characteristics has not been established. To provide a direct comparison of the two algorithms, we used the GRACE2.0 features and re-fitted the GRACE2.0 using our training data. The re-fitted GRACE2.0 score had the same model discrimination and better model calibration than the original version (Fig. 2), but still had inferior prediction compared to PMHnet.

The above observations are in good agreement with the current literature on ML-based secondary risk-stratification models, which have found machine learning models to offer better performance than simpler, existing scores in current clinical use^10–13, 40, 41^. A transition towards feature-rich models that utilize more of the available data can therefore be an advantage. The 584 features used in our final model are neither bespoke nor specifically collected for this decision-support application, and instead represent clinical information gathered during routine work-up, management and treatment of patients. Using models that rely on several hundred features means that the current practice of manually entering data into a webtool becomes very impractical^42^. Instead, novel risk-prediction models need to be fully integrated in the EHR systems such that data can be automatically pulled and integrated.

Among previously published machine-learning models for secondary prediction in IHD, our study has several strengths. Firstly, whereas almost all the existing literature uses binary classification, the use of survival or time-to-event models is less explored. One notable exception is the model reported by Steele et al.^10^ which employed random survival forests and elastic net Cox regression to predict mortality in patients (n=80,000) with a history of coronary artery disease. One of the defining characteristics of survival models is the ability to handle censored data^43^, which in other types of models would have been left out. In addition, survival models can distinguish between “died after a week” and “died after 10 months” which e.g., would be identical in a 1-year binary classification model. To the best of our knowledge, we are the first to use neural network-based survival models in this context. Secondly, we externally validated our model using data from a different country. Most models in the literature were not suboptimal externally validated^10, 11, 40, 41^, and among those that have been, only two used data from a different country^12, 13^. Demonstrating that a given model can accurately predict beyond borders is crucial for generalizability. Thirdly, our model can predict all-cause mortality up to five years after the index date. Probably owing to the fact that survival models have not been used, most models in the domain exclusively operate with a prediction horizon of one or two years^12, 13, 40, 41^. Longer prediction horizons might be necessary for long-term disease management.

Although the features we have used represent data collected from a typical clinical workflow and therefore should be generally applicable, inter-regional and inter-national differences in clinical practice may affect what data is available and when. This is for instance exemplified by differences in diagnostic work-up, or timely access to coronary angiography, but may also relate to differences in access to previous medical data. Such aspects could affect the generalizability of our included features, but with internationally accepted treatment guidelines the differences should be minimal. Possible solutions are to reduce slightly the complexity of models to better match the intersection of features available across countries/regions and/or re-fitting and possibly retraining the model each time it is deployed in a new setting. In our external validation we used data from Iceland, which in an international perspective is very similar to Denmark when comparing healthcare systems^44, 45^. For that reason, we downscaled the model slightly to account for data availability, but re-training on Icelandic data was not deemed necessary. That being said, using the Icelandic data, we found evidence of miscalibration as the model was found to overestimate the risk for some patients and this suggests that further adjustment of the model is needed were it to be deployed in Iceland. However, since miscalibration does not affect the accurate risk-stratification of patients, we did not pursue that issue further in this study. For future studies, we note that techniques such as Platt-scaling or isotonic regression could be used to remedy calibration-issues^46^.

The dynamic nature of clinical environments can lead to deterioration of model performance^47, 48^. Advances in treatment and diagnosis mean that the baseline risk of patients with ischemic heart disease could change over time, which in turn would lead to a drift of model calibration. As an example, the logistic EuroSCORE^49^, a pan-European risk-stratification model for cardiac surgery published in 2003 was since its inception gradually found to overestimate mortality^50^. The EuroSCORE has since been replaced by EuroSCORE II which for the time being has remedied these issues^51^. This type of systematic decline of model performance has important implications for our model as well and necessitates that the model performance is continuously monitored to ensure acceptable up-to-date performance. The need for real-time monitoring of model performance is another strong argument for risk-stratification tools to be tightly integrated within EHR systems.

As a secondary analysis in this study, we tested adding a panel of polygenic risk scores to the input features in PMHnet and assessed how they might impact model performance. The 19 different genetic risk scores were included based on being related to cardiometabolic health and covered traits such as *blood pressure*, and *total cholesterol*, but also included *heart failure* and *acute myocardial infarction*. Where for example the *acute myocardial infarction* risk score is developed for primary risk prediction, i.e., disease development, our model is concerned with secondary risk prediction, i.e., modelling risk for those who already have the disease. It is known that CAD PRS associate with events in both primary and secondary event populations, however, these PRSs are PRSs of prevalent diseases, not mortality. Whether or not a primary risk score is useful in a secondary risk context is clear upfront, but as parts of the underlying disease process are known to be shared between the two, we hypothesized that the score could be used in our context. Focusing the analysis only on the subset of patients for which we could obtain genotypes (n=13,449, 37.4%), we found no significant difference in performance after adding the PRS scores to either *model-1* (*Diagnoses*) and *model-2* (*Diagnoses* + *ClinicalOne*). As the 585 features include the prior disease history recorded over more than 25 years, our interpretation is that a “realized” life-course disease trajectory is more informative than the germline risk that can be calculated from the genotype. Moreover, the disease trajectory also holds information on life-course exposures and may therefore also include exposure information that quite implicitly is related to genetic data only. The version of PMHnet trained on the disease history only, with and without genetics, indicates that there is little gain from the PRS tested in this case. As we did not have genetic data for the full cohort, we cannot exclude that our interpretation is affected by this aspect.

Prospective studies are needed to ascertain how feature-rich risk-stratification methods can be used to alter, guide, and hopefully improve treatment. The ability to accurately predict high-risk is useful for identifying patients that may benefit from more extensive treatment and more frequent visits at the hospital. In contrast, accurate identification of low-risk patients may potentially be used to limit work-up, extent of pharmacological treatment and follow-up content and intensity and thereby prevent potential harmful overtreatment. Striking the correct balance between over- and undertreatment can contribute to the advancement of precision medicine, and here a well-calibrated and highly discriminative risk-prediction model can serve as an important tool.

In routine cardiology, a multitude of diagnostic, prognostic, and treatment-related scores are applied. However, all the presently applied scores are based on a very limited number of features. The present findings indicate a significantly added value of applying far more features and of introducing machine-learning. Thus, precision treatments in cardiology may benefit from using more features and machine-learning to replace the present scores.

## Data Availability

Due to national and EU regulations, the datasets used for model development and validation cannot be made publicly available. Research groups with access to secure and dedicated computing environments can request access to the source data registries via application to the Danish Health Data Authority.

## Acknowledgements

We acknowledge Mette Hartlev, Franziska Walder, Mette Gørtz, and Katharina Ó Cathaoir for helpful comments and discussions in the writing of this manuscript.

